# Laboratory Testing Implications of Risk-Stratification and Management for Improving Clinical Outcomes of COVID-19 Patients

**DOI:** 10.1101/2020.06.12.20129098

**Authors:** Caidong Liu, Ziyu Wang, Jie Li, Changgang Xiang, Lingxiang Wu, Wei Wu, Weiye Hou, Huiling Sun, Youli Wang, Zhenling Nie, Yingdong Gao, Ruisheng Zhang, Xinyi Xia, Qianghu Wang, Shukui Wang

## Abstract

The high mortality rate of COVID-19 patients is mainly caused by the progression from mild to critical illness. To identify the key laboratory indicators and stratify high-risk COVID-19 patients with progression to severe/critical illness, we compared 474 moderate patients and 74 severe/critical patients. The laboratory indicators, including lactate dehydrogenase (LDH), monocytes percentage, etc. were significantly higher in the severe/critical patients (P <0.001) and showed a noticeable change at about a week before the diagnosis. Based on these indicators, we constructed a risk-stratification model, which can accurately grade the severity of patients with COVID-19 (accuracy = 0.96, 95% CI: 0.94 - 0.989, sensitivity = 0.98, specificity = 0.84). Also, compared with non-COVID-19 viral pneumonia, we found that COVID-19 had weaker dysfunction to the heart, liver, and kidney. The prognostic model based on laboratory indicators could help to diagnose, monitor, and predict severity at an early stage to those patients with COVID-19.

## INTRODUCTION

The global outbreak of COVID-19 highlights the importance of early and rapid diagnosis, monitoring, risk assessment, and medical resource management in the prevention and control of epidemics. The high mortality rate of COVID-19 patients is mainly caused by the progression from the mild condition to the critical illness^1^. Therefore, it is an urgent need for effective methods to predict prognosis early. At present, nucleic acid detection and antibody detection are the main technical approaches for clinical diagnosis of COVID-19 patients but fail to help to judge whether a patient will progress to severe or critical illness ^2, 3, 4^. Besides, CT imaging lacks specificity, requires a large number of professional technicians, and easily exhausts resources when the epidemic is serious. The latest research shows that, based on artificial intelligence methods, CT can be used to diagnose COVID-19 quickly. However, the accuracy of using CT alone to predict patient severity is only about 80%^5^.

Previous studies have reported that in the early published 41 COVID-19 cases, five patients presented with varying degrees of myocardial injury, cardiovascular disease patients are more likely to develop into critical patients after COVID-19 infection, and the risk of death is higher^6^. The abnormal of different laboratory indicators can represent damage to different organs. For example, NT-proBNP indicates cardiac dysfunction and Alkaline phosphatase (ALP) indicates liver dysfunction. In addition, other laboratory indicators are highly correlated with the risk of disease progression, such as the LYM, IL-6, etc.^7^. These findings suggested that the laboratory indicators can be used to predict the severity of COVID-19 pneumonia patients. Therefore, this study aims to analyze the laboratory indicators associated with a severity that can build a risk-stratification model for scientific screening of COVID-19 critical patients.

## RESULTS

### Clinical characteristics

We reported 548 cases enrolled from the First People’s Hospital of Jiangxia District of Wuhan, of which 205 patients had complete clinical information, including 57 severe or critical (severe/critical) patients, and 148 moderate COVID-19 patients. The average age of these patients was 52.4. Notably, the median age of severe/critical patients was significantly higher than that of moderate patients (p<0.01, **Table 1**). There was no significant difference in disease susceptibility between males and females (p=0.84, **Table 1**). Besides, the comorbidities, such as hypertension, diabetes, cerebrovascular disease and cardiovascular disease, were significantly associated with severe/critical patients (p<0.01, **Table 1**). There was no significant difference in other indicators, such as symptoms, pulse, and respiratory rate between the two groups (**Table 1**).

**Table 1.** Clinical characteristics of 148 non-critical and 57 critical/severe patients.

|  | Total<br>(n=205) | Non-critical<br>(n=148) | Critical/Severe<br>(n=57) | P value |
| --- | --- | --- | --- | --- |
| Age (x±s) | 52.4±14.2 | 49.9±13.2 | 59.1±14.3 | <0.01 |
| Gender |  |  |  |  |
| Male (n, %) | 103 (50.2) | 75 (50.7) | 28 (49.1) | 0.84 |
| Female (n, %) | 102 (49.8) | 73 (49.3) | 29 (50.9) | 0.84 |
| Wuhan residence history or close contact with confirmed cases (n, %) | 69 (33.7) | 51 (34.5) | 18 (31.6) | 0.70 |
| Comorbidities |  |  |  |  |
| COPD | 2 (1.0) | 1 (0.7) | 1 (1.8) | 0.48 |
| Diabetes (n, %) | 15 (7.3) | 6 (4.1) | 9 (15.8) | 0.004 |
| Hypertension (n, %) | 22 (10.7) | 7 (4.7) | 15 (26.3) | <0.001 |
| Chronic liver disease (n, %) | 10 (4.9) | 6 (4.1) | 4 (7.0) | 0.37 |
| Chronic kidney disease (n, %) | 9 (4.4) | 4 (2.7) | 5 (8.8) | 0.057 |
| Cerebrovascular disease (n, %) | 12 (5.9) | 1 (0.7) | 11 (19.3) | <0.001 |
| Cardiovascular disease (n, %) | 11 (5.4) | 4 (2.7) | 7 (12.3) | 0.006 |
| Malignancy (n, %) | 3 (1.5) | 1 (0.7) | 2 (3.5) | 0.13 |
| Symptoms |  |  |  |  |
| Dry cough (n, %) | 162 (79.0) | 117 (79.1) | 45 (78.9) | 0.99 |
| Diarrhea (n, %) | 13 (6.3) | 8 (5.4) | 5 (8.8) | 0.38 |
| Fever (n, %) | 123 (60.0) | 85 (57.4) | 38 (66.7) | 0.23 |
| Dyspnea (n, %) | 14 (6.8) | 9 (6.1) | 5 (8.8) | 0.49 |
| Expectoration | 25 (12.2) | 16 (10.8) | 9 (15.8) | 0.33 |
| Myalgia | 61 (29.8) | 47 (31.8) | 15 (26.3) | 0.45 |
| Nausea | 18 (8.8) | 12 (8.1) | 6 (10.5) | 0.58 |
| Headache | 12 (5.9) | 9 (6.1) | 3 (5.3) | 0.82 |
| Vomiting | 8 (3.9) | 4 (2.7) | 4 (7.0) | 0.15 |
| Fatigue | 102 (49.8) | 73 (49.3) | 29 (50.9) | 0.84 |
| Signs |  |  |  |  |
| Pluse,median (IQR) | 86 (79~93) | 87 (81~93) | 84 (75~94) | 0.12 |
| Systolic pressure (x±s), mmHg | 125.6±11.3 | 122.3±9.7 | 129.7±12.4 | <0.01 |
| Diastolic pressure (x±s), mmHg | 77.9±10.2 | 75.6±9.4 | 81.5±10.9 | <0.01 |
| Respiratory rate,median (IQR) | 20 (19~21) | 20 (19~22) | 20 (19~20) | 0.35 |

### SARS-CoV-2 produces less dysfunction than other viral pneumonia

Increasing studies showed that the infection of viral pneumonia might be associated with organ dysfunction^8, 9, 10, 11^. We hence explored the change of organ function-related indicators during hospitalization for 548 COVID-19 cases and eighteen other viral pneumonia (designated as non-COVID-19) cases. Interestingly, we found that there were thirteen indicators showing significant differences between the two groups (**Table 2**, Wilcoxon two-sided rank-sum test, p<0.05). Most of these laboratory indicators were related to abnormal heart, liver, and kidney functions which may be due to cytokine storm caused by pneumonia ^12^. N-terminal pro-brain natriuretic peptide (NT-proBNP) is an important indicator ^13^, reflecting heart function. In recent years, many studies have found that inflammatory factors can also stimulate the increase of serum NT-proBNP levels. Our studies showed that patients in non-COVID-19 group had higher levels of NT-proBNP than those of COVID-19 group (1259.4pg / mL vs. 90.285pg / mL, p=0.045), suggesting these viral pneumonia might lead to more severe heart damage. Besides, the level of LDH in non-COVID-19 was higher than COVID-19 patients (594 vs. 242.85, p<0.001), indicating multiple potential causes, including acute kidney disease, acute liver disease. The level of alanine aminotransferase (19 U / L vs. 40U / L, p <0.001) and aspartate aminotransferase (30.1s vs. 36s, p <0.001) were higher in non-COVID-19 group. The median activated partial thromboplastin time was longer than that in the COVID-19 group. The median level of albumin and hemoglobin decreased by more than 5g/L and 10 g/L in non-COVID-19 patients, respectively (albumin: 33.8 g / L vs. 38.95 g / L, p<0.001; hemoglobin: 121 g / L vs. 135.25 g / L, p=0.003). Hypoalbuminemia is frequently observed in conditions like diabetes, hypertension, and chronic heart failure^14^. In summary, these findings demonstrated that most patients with COVID-19 pneumonia produce less dysfunction and toxicity.

**Table 2.** Laboratory findings of patients between COVID-19 and Non-COVID-19.

|  | Normal<br>Range | Total<br>(N=403) | Non-COVID-19<br>(N=18) | COVID-19<br>(N=385) | P-value |
| --- | --- | --- | --- | --- | --- |
| **Laboratory Findings** |  |  |  |  |  |
| **Infection markers** |  |  |  |  |  |
| Eosinophil<br>percentage, % | 0.4~8 | 0.54(0.2-1.3) | 2(0.1-3.5) | 0.5(0.2-1.2) | 0.035 |
| White blood cell<br>count, g/L | 3.5~9.5 | 5.7(4.4-7.4) | 4.42(2.3-7.2) | 5.7(4.6-7.4) | 0.005 |
| **Liver injury markers** |  |  |  |  |  |
| Albumin, g/L | 35~55 | 38.5(35-42) | 33.8(30-36) | 38.95(36-42) | <0.001 |
| Alanine transaminase | 9~50 | 20.1(13-34) | 40(23-96) | 19(12-29) | <0.001 |
| **Heart injury markers** |  |  |  |  |  |
| Aspartate<br>Transaminase | 13~35 | 29.6(21-45) | 51(30-82) | 27.4(20-41) | <0.001 |
| Lactate<br>Dehydrogenase, U/L | 80~285 | 259.15(210-380) | 594(470-830) | 242.85(200-330) | <0.001 |
| N-terminal pro-brain<br>natriuretic peptide | 0~125 | 94.415(51-440) | 1259.4(340-2300) | 90.285(49-410) | 0.045 |
| **Blood examination** |  |  |  |  |  |
| Activated partial<br>thromboplastin time | 24~36 | 30.25(28-32) | 36(35-42) | 30.1(28-32) | <0.001 |
| Hematocrit, % | 40~50 | 39.8(36-43) | 36.775(26-40) | 39.85(36-43) | <0.001 |
| Hemoglobin, g/L | 130~175 | 135(120-150) | 121(95-130) | 135.25(120-150) | 0.003 |
| Calcium, mmol/L | 2.08~2.8 | 2.09(2-2.2) | 1.97(1.9-2) | 2.1(2-2.2) | <0.001 |
| Potassium, mmol/L | 3.5~5.3 | 3.95(3.6-4.3) | 3.405(3.2-3.8) | 4.02(3.7-4.4) | <0.001 |
| Sodium, mmol/L | 137~147 | 139(136-141) | 137.8(135.2-139.5) | 139(136-141) | 0.032 |

### The patients with heart disease, hypertension, or cancer were more likely to develop into severe/critical COVID-19

We next explored the impact of the underlying diseases on the progression of COVID-19. Based on the analysis of 548 COVID-19 cases, we found only 9% of patients without underlying disease progressed to severe or critical condition. In contrast, 16% of severe/critical patients were diagnosed with at least one underlying disease (**Figure 1A**), suggesting that COVID-19 patients with comorbidities were prone to develop severe or critical illness. Furthermore, we extended the analysis to 3015 COVID-19 cases enrolled from Huoshenshan hospital, in Wuhan, China, of which 1452 and 1563 patients were classified as moderate and severe/critical COVID-19.

**Figure 1.**
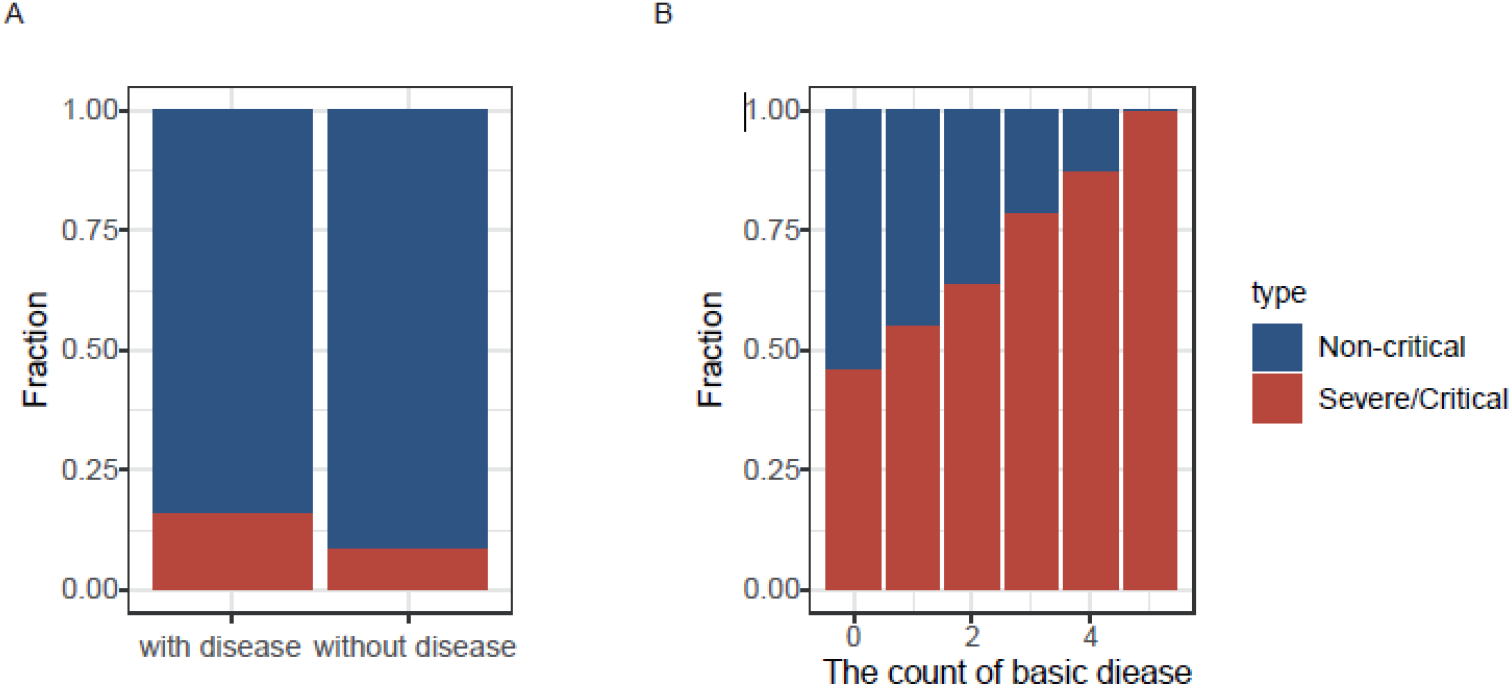
The impact of the underlying disease on the progression of COVID-19. **A**. Distribution of critical patients in different numbers of underlying diseases in 548 COVID-19 patients. **B**. Distribution of critical patients in different numbers of underlying diseases in 3015 COVID-19 patients.

Among these cases, 565 out of 1563 (36.1%) severe/critical patients had hypertension, which was 1.5-fold higher than that of moderate patients (24.5%) (Fisher’s exact test, p<0.0001, odd ratio=1.74). 409 patients had diabetes, and 61% progressed to severe/critical COVID-19 (Fisher’s exact test, p=0.0001, odd ratio=1.5). 240 patients suffered from heart disease and 170 patients developed to severe/critical COVID-19 (Fisher’s exact test, p < 0.0001, odd ratio=2.4). 74% patients with cancer were diagnosed as severe/critical COVID-19 (Fisher’s exact test, p=0.0001, odd ratio=2.7). On the contrary, kidney (p=0.07) and liver (p=0.8) disease had no association with the progression of COVID-19.

Moreover, with the increase of underlying diseases, the proportion of severe/critical patients was prone to increase (**Figure 1B**). 31 of 39 cases with hypertension, diabetes, and heart disease were diagnosed as severe/critical COVID-19. Increasing studies showed that the infection of COVID-19 may be associated with the high expression of ACE2. It is reported that the expression of ACE2 is substantially increased in patients with diabetes, heart disease, and hypertension^15, 16^. Finally, logistic regression was performed for the underlying disease. Hypertension (coefficient 95% CI: 1.3-1.8, p<0.001), heart disease (coefficient 95%CI: 1.5-2.7, p<0.001), and cancer (coefficient 95% CI: 1.4-4.1, p=0.0015) were significantly associated with the severity of COVID-19.

### Identification of laboratory indicators related to the severity of COVID-19

We next explored the difference of laboratory findings between moderate and severe/critical COVID-19 cases. After excluding 163 cases without routine indicators before diagnosis, we focused on the analysis of 385 cases consisting of 329 moderate and 56 severe/critical cases.

We found the high risk factors related to the progression of COVID-19 included procalcitonin (PCT), C-reactive protein (CRP), neutrophils percentage, lymphocytes percentage, LDH, NT-proBNP, and high-sensitivity troponin T (hs-cTnT) (Wilcoxon rank-sum test, p<0.001, **Table 3**), etc. We noted that most of the severe/critical patients presented lymphopenia and elevated levels of inflammatory biomarkers. The levels of PCT in severe/critical patients at the initial stage were higher than those in moderate patients (0.225 vs. 0.06, p<0.001), suggesting serial procalcitonin measurement may play a role in predicting evolution towards a more critical form of the disease^17^. The CRP showed a similar trend to PCT, which became significantly higher in severe/critical patients (44.5 vs. 21.8, p<0.001). Lymphocyte percentage was significantly higher in the moderate COVID-19 patients than severe/critical COVID-19 patients (22.4% vs 13.8%, p<0.001), which was consistent with the finding reported by Zhou et al., where lymphocyte count was lower in non-survivors than survivors ^7^. The percentage of neutrophils was elevated along with the severity of COVID-19 (77.8 vs. 66.4, p<0.001). Besides, NT-proBNP (292.1 vs. 80.34, p=0.0021), hs-cTnT (0.016 vs. 0.008, p<0.001), and LDH (314 vs. 235, p<0.001) of severe critical patients were significantly higher than those of moderate patients. LDH was a risk factor associated with disease progression in patients infected with 2019-nCoV^18^. Many types of research proved that elevated NT-proBNP was significantly correlated with critical disease^19^. The clinical indicators between mild and severe patients showed a noticeable change at about a week before the diagnosis (**supplementary Figure 1**).

**Table 3.**
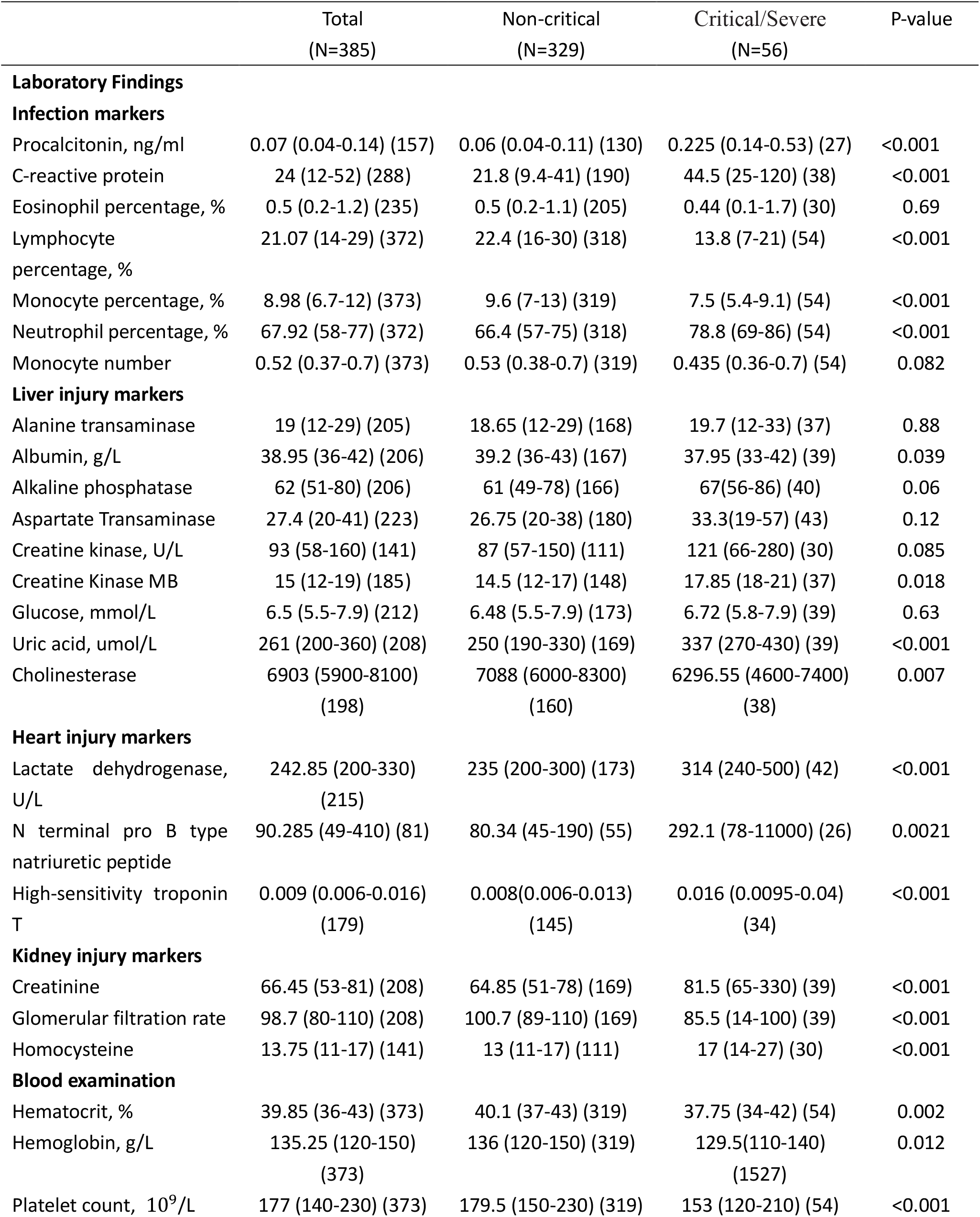
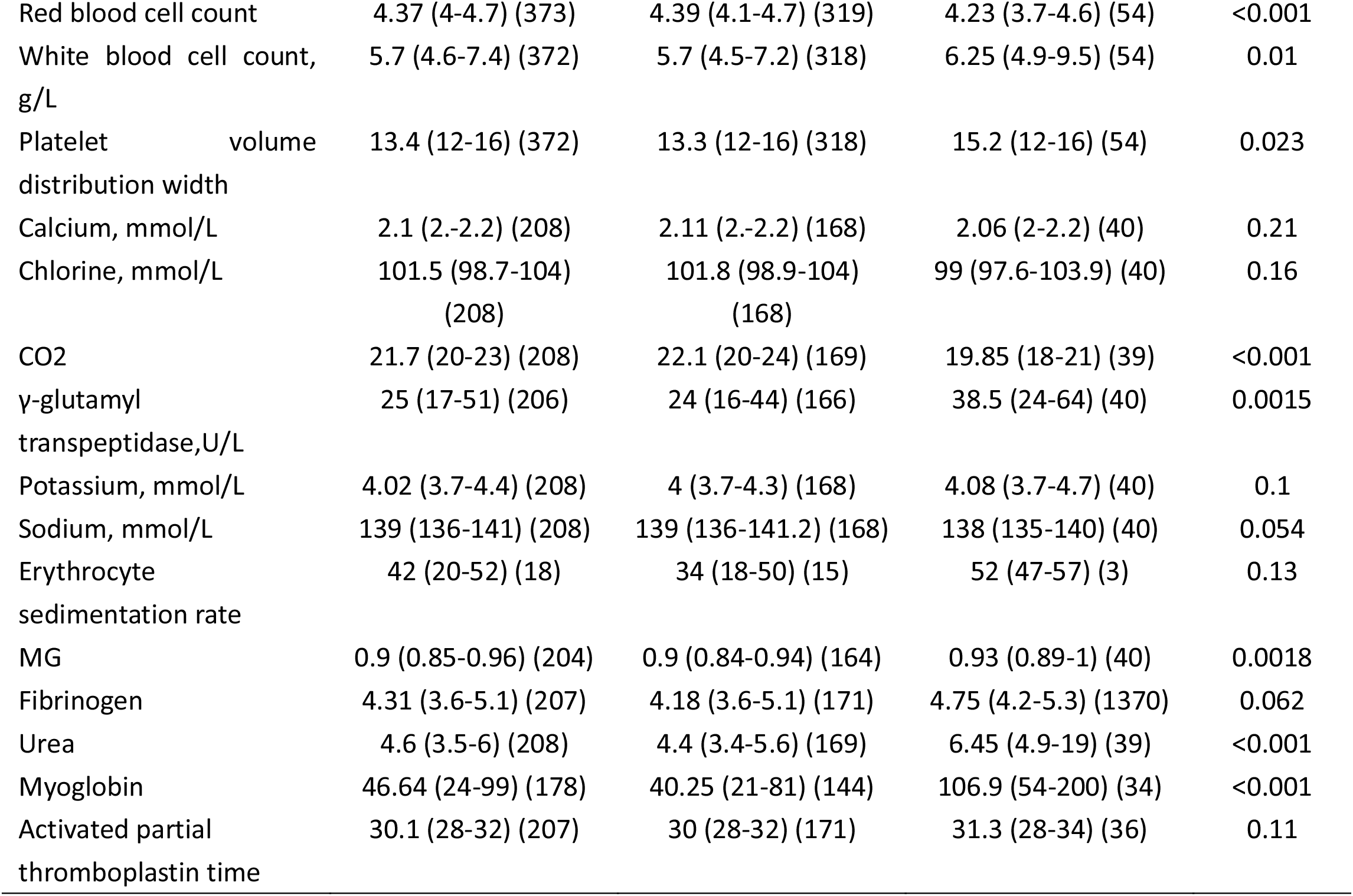
Laboratory findings of patients between non-critical and critical/severe.

In summary, most of the severe/critical patients presented the increase of neutrophil percentage, procalcitonin, and the decrease of lymphocyte percentage, which may potentially induce a cytokine storm in the body, and generate a series of immune responses to damage the corresponding organs^12, 20^.

### A prognostic model for evaluating the severity of COVID-19 patients by laboratory indicators

To validate that whether laboratory findings could predict the progression of COVID-19, we selected laboratory indicators with less than 80% missing values and performed principal component analysis (PCA) on the entire patient cohort, including 325 moderate cases and 56 severe/critical cases. Four cases with more than 80% missing values were excluded. The result showed that there was an essential difference in laboratory indicators between moderate and severe/critical patients, including NT-proBNP, LDH, neutrophils, and LYM, etc., suggesting that these inidcators may play an important role in the principal component analysis (**Figure 2A**).

**Figure 2.**
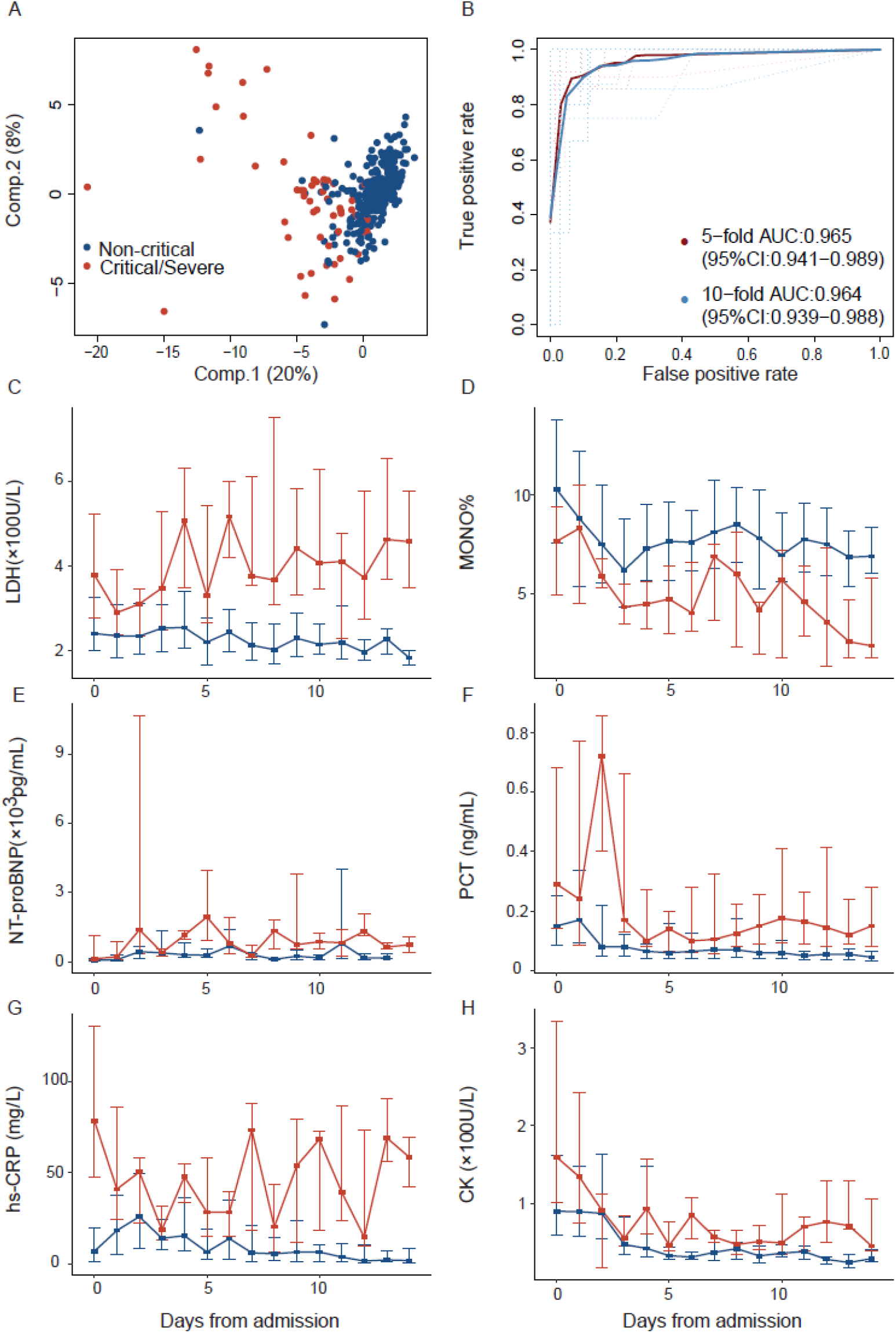
Laboratory findings for predict progression of COVID19. **A**: PCA based on laboratory findings. The red represents the severe patients and blue represent the mild patients. **B**: The ability of the model to distinguish between severe and mild patients. the x-axis is specificity and the y-axis represents sensitivity. The red represents the five-fold cross-validation and blue represents the five-fold cross-validation. **C-H**: The change of clinical markers during the period of admission. The red represents the severe patients and blue represent the mild patients. The dots represent the median, and the horizontal lines represent the upper and lower quartiles.

We next applied these indicators to ensemble learning-based classifier, followed by five-fold cross-validations, 10-fold cross-validation, and leave-one-out validation. The representative receiver operating characteristic (ROC) for the entire cohort was shown in **Figure 2B**. The average sensitivity and specificity of five cross-validations were 0.95 and 0.92, respectively. The sensitivity ranged from 0.86 to 1, and specificity ranged from 0.84 to 0.97. The average AUC of the five cross-validations was 0.96 (AUC 95% CI:0.941-0.989). It still achieved satisfying results in ten-fold cross-validation (mean sensitivity and specificity, 0.94 and 0.96, respectively, AUC 95% CI:0.939-0.988). The accuracy of leave-one-out method was 0.96 (specificity:0.98, sensitivity:0.84). In the latest research, Kang Zhang, etc. have developed an artificial intelligence (AI) tool, which has the functions of severity classification and critical illness prediction. However, it uses chest CT images to predict the development of patients with critical illness with an AUC of 0.85. At the same time, the AUC predicted by other laboratory indicators can be increased to 0.91. This shows that the accuracy of the system in predicting the severity of patients is lower than our prediction model using laboratory indicators alone^5^.

We finally paid attention to the features that appeared at high frequencies in both 5-fold cross-validation and 10-fold cross-validation, including the PCT, LDH, and NT-proBNP, etc. All of these indicators remained relatively stable along with the time from admission to diagnosis. However, the disparity of indictor levels between moderate and severe/critical patients was prone to widen (Figure1C-1G). LDH was higher in the severe/critical patients and escalated with the progression of the disease (**Figure 1C**). The trend of monocyte was completely opposite (**Figure 1D**). Besides, we found that there was no significant difference in clinical indicators between patients without underlying disease and patients with underlying disease (**supplementary Figure 2**). These results suggest that the application of laboratory findings to predict the progression of COVID-19 patients can act as a useful complement to conventional methods.

## DISCUSSION

The severe/critical COVID-19 patients maintained a high mortality rate due to a lack of efficient therapeutic approaches during the pandemic. It is an urgent need for effective methods to predict and monitor the progression of COVID-19 patients from moderate to severe or critical conditions. Based on the 548 COVID-19 cases enrolled from in the First People’s Hospital of Jiangxia District of Wuhan, China, we systematically explored the difference of comorbidity rate and laboratory findings. In our study, we found that patients with comorbidity were prone to develop into severe or critical illness. The proportion of severe/critical condition positively associated with the increase in the number of comorbidities diagnosed in the patients. Notably, the analysis based on the logistic regression model showed that patients with heart disease, hypertension, or cancer were more likely to be suffering from the progression of COVID-19. Many studies demonstrated that these underlying diseases might promote the expression of ACE2^15, 16^, leading to a high-risk of COVID-19 infection.

We found the high-risk factors related to the progression of COVID-19 included PCT, CRP, neutrophils and lymphocytes percentage, LDH, NT-proBNP, and hs-cTnT, etc. Most of these laboratory indicators were reported to be associated with the cytokine storm, suggesting a series of immune responses to damage the corresponding organs. Based on these indicators, we finally constructed a risk-stratification model by using an ensemble learning-based classifier, which presented an AUC of more than 95%. Besides, by exploring the change of organ function-related indicators during hospitalization for COVID-19 and non-COVID-19 cases. We found that the patients in the COVID-19 group were diagnosed with less dysfunction and toxicity, suggesting that SARS-CoV-2. We anticipate that our study will provide vital information for helping clinicians to diagnose and monitor the progression of COVID-19 patients.

## METHODS

### Inclusion criteria and clinical classification

Diagnostic criteria: The diagnosis of COVID-19 is based on the “New Coronavirus Pneumonia Diagnosis and Treatment Plan (provisional 6^th^ Edition)” issued by the National Health and Health Commission. It should meet the diagnostic conditions of suspected cases, meanwhile, the quantitative reverse transcription polymerase chain reaction (qRT-PCR) detection of specimens such as sputum, throat swabs, lower respiratory tract secretions should be positive for SARS-CoV-2 nucleic acid (http://www.kankyokansen.org). Data inclusion criteria: At the same time that the above diagnostic criteria were met, the time between the reporting of laboratory test results and the patient’s admission was less than 24h. Clinical classification: According to the disease classification criteria of the “New Coronavirus Pneumonia Diagnosis and Treatment Scheme (Trial Version 6)”, symptoms such as fever and respiratory tract and pulmonary imaging manifestations were diagnosed as mild/moderate (Non-critical) type, one of the following conditions was diagnosed as severe (critical) type: (1) shortness of breath, respiratory rate ≥ 30 times/min; (2) oxygen saturation ≤ 93% in resting state; (3) arterial partial pressure of oxygen (PaO2) / oxygen concentration ≤ 300mmHg; (4) pulmonary imaging showed that the lesions progressed more than 50% within 24-48h.

### Data collection

From December 1, 2019 to February 13, 2020, a total of 385 cases of confirmed COVID-19 patients meeting the above inclusion criteria were collected through the hospital electronic medical record system. The collected information includes: the time of patient admission, clinical diagnosis, condition evaluation, test report time and and laboratory test information, etc. This study was approved by the hospital ethics committee.

### prognostic model

To distinguish moderate COVID-19 patients from severe/critical COVID-19 patients, we constructed a random forest classifier based on adaboost using the training cohort of 385 patients. The R adabag package was then used to perform the ensemble learning. Prediction accuracy was determined from a randomly 25% sample of the training data as well as the independent test set. Because many diverse factors are associated with different disease states, the feature for classification was drawn from the blood routine examination, blood biochemical examination, and laboratory findings. All indicators are checked before the patient is diagnosed. Multiple measurements of the same patient are replaced by the mean value. Missing values are completed by conditional mean completer.

### Statistical Analysis

The median (IQR) and n (%) are used to show continuous variables, and we used the wilcoxon rank sum test to compare the differences of continuous variables between groups. In the bilateral test, the index of p <0.05 is considered statistically significant. All statistical analyses were performed using R software.

## Data Availability

The datasets generated during and/or analyzed during the current study are available from the corresponding author on reasonable request.

## Author Affiliations

Department of laboratory medicine (S.W., C.L., C.X., W.H., Y.W., Z.N., R.Z., Y.G.), and the General clinical research center (H.S., H.W., K.Z., X.X.), Nanjing First Hospital, Nanjing Medical University, Nanjing 210006, Jiangsu, China; Department of Bioinformatics (Q.W., Z.W., L.W., W.W.), and the Jiangsu Key Lab of Cancer Biomarkers, Prevention and Treatment, Collaborative Innovation Center for Personalized Cancer Medicine (Q.W.), Nanjing Medical University, Nanjing, Jiangsu 211166, China; Collaborative Innovation Center for Cardiovascular Disease Translational Medicine, Nanjing, Jiangsu 211166, China (Q.W.); COVID-19 Research Center, Institute of Laboratory Medicine, Jinling Hospital, Nanjing University School of Medicine, Nanjing, Jiangsu 210002, China (X.X.); Department of Laboratory Medicine & Blood Transfusion, Wuhan Huoshenshan Hospital, Wuhan, Hubei 430100, China (X.X.); Joint Expert Group for COVID-19, Wuhan Huoshenshan Hospital, Wuhan, Hubei 430100, China (X.X.).

## Author Contributions

Wang Shukui, Qianghu Wang, Xinyi Xia had full access to all of the data in the study and takes responsibility for the integrity of the data and the accuracy of the data analysis. Caidong Liu, Ziyu Wang, Jie Li, and Changgang Xiang contributed equally. Concept and design: Wang Shukui, Qianghu Wang. Data collection: Caidong Liu, Changgang Xiang, Weiye Hou, Huiling Sun, Haoyu Wang, Kaixuan Zeng, Xueni Xu, Youli Wang, Zhenling Nie, Ruisheng Zhang, Yingdong Gao. Data analysis and interpretation: Lingxiang Wu, Ziyu Wang, Wei Wu, Jie Li.

---

**Supplementary Figure 1:**
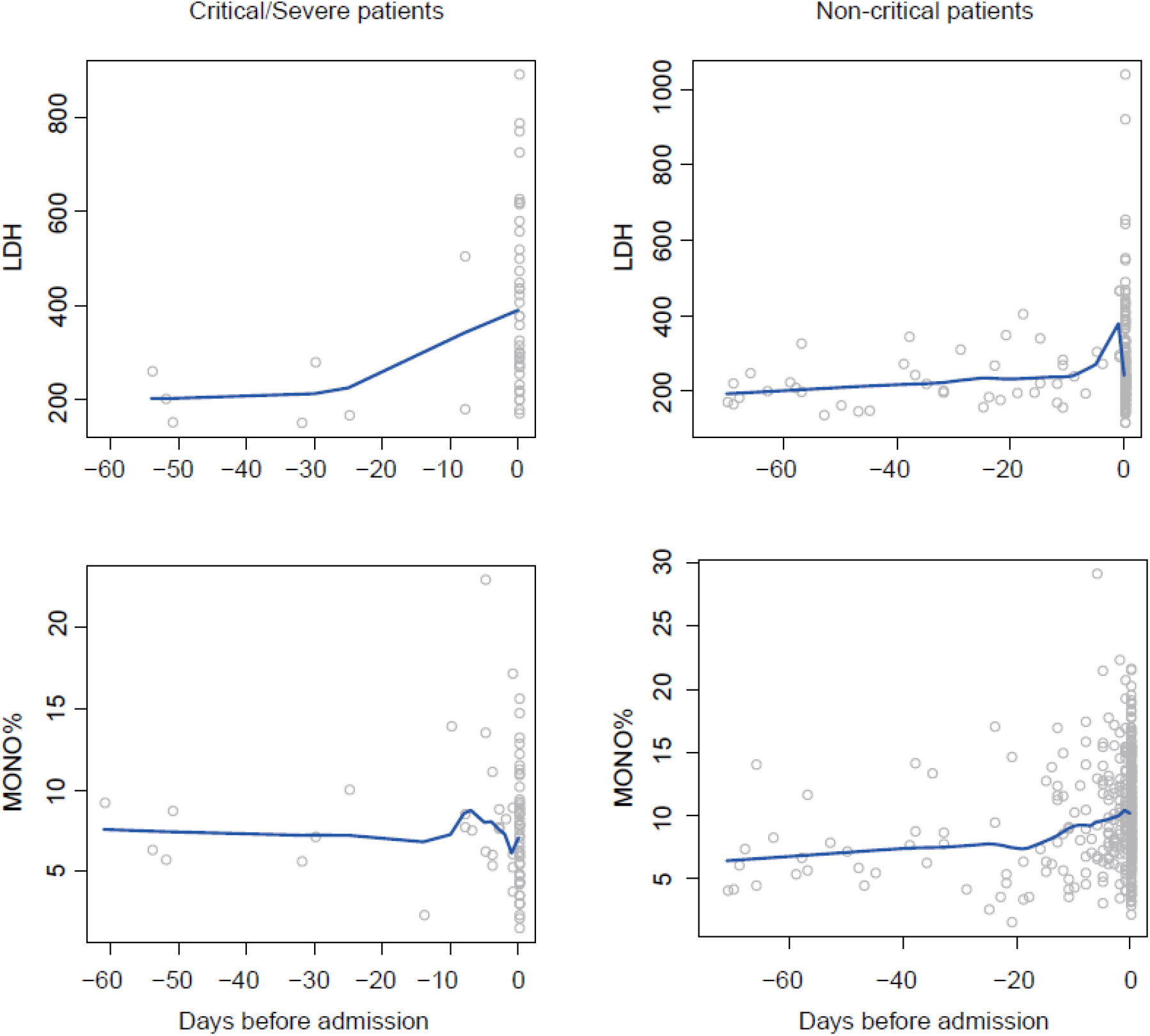
The change of clinical markers during the period before admission. The blue line is the fitting curve.

**Supplementary Figure 2:**
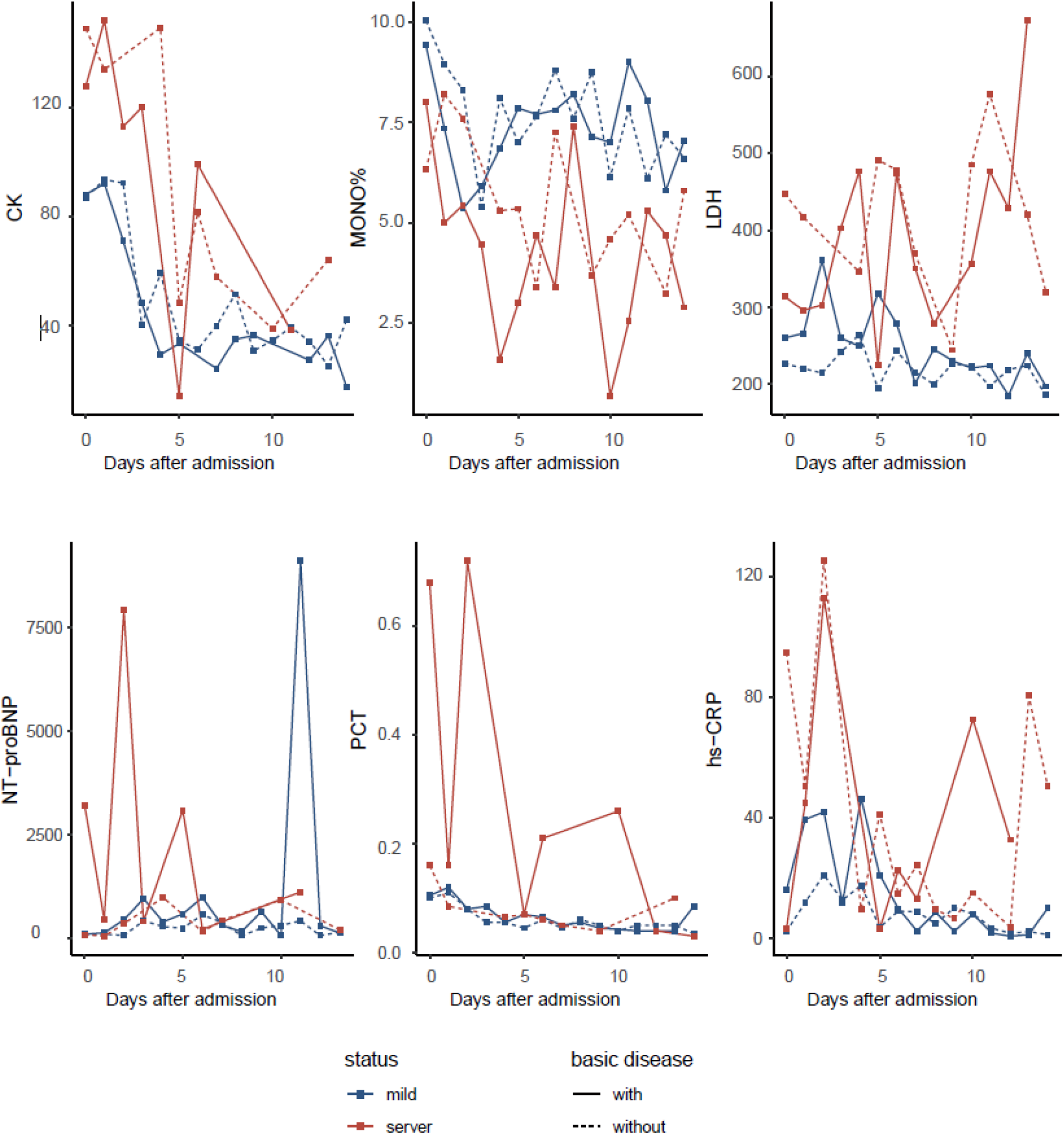
The change of clinical markers during the period of admission. The red represents the severe patients and blue represent the mild patients. The dots represent the median, and the horizontal lines represent the upper and lower quartiles. The solid line represents the patients with underlying disease, and the dotted line represents the patients without underlying disease.

